# Implementation of a Gait and Balance Battery of Outcome Measures in an Inpatient Rehabilitation Hospital

**DOI:** 10.1101/2025.11.17.25340425

**Authors:** Leah Ling, Randy Carson, Geneviève N. Olivier, Margaret A. French

## Abstract

**Importance:** Standardized outcome measure (OM) batteries are important for tracking functional progress, enhancing communication among providers, and evaluating intervention effectiveness in physical therapy. However, implementation of standardized batteries is challenging due to time constraints, documentation requirements, and limited resources.

**Objective:** To evaluate implementation of a standardized gait and balance OM battery by physical therapists for patients with stroke, brain injury, and spinal cord injury in the inpatient rehabilitation facility (IRF) setting.

**Design:** A retrospective cohort study of 773 episodes of care at the IRF, and a quantitative survey for 22 staff physical therapists at the IRF.

**Setting:** A 75-bed IRF in the United States.

**Interventions:** A standardized OM battery with five measures was implemented by all physical therapy teams at the IRF.

**Main Outcomes and Measures:** We assessed four implementation outcomes: Penetration and Sustainability, which we assessed with retrospective cohort data of episodes of care, and Acceptability and Feasibility, which we assessed with the quantitative survey of physical therapists.

**Results:** The implementation effort resulted in a significant improvement in the completion rates of all five OMs, with most measures exceeding a 70% completion rate threshold within one year after implementation. Completion rates of the entire battery increased significantly with the implementation effort, though did not reach the goal 70% completion rate. Physical therapists indicated that implementation of the standardized OM battery resulted in positive practice changes, and that barriers to completion were patients’ functional levels and the time required for completion.

**Conclusions:** Implementation of a standardized OM battery is feasible in the IRF setting, though OM completion rates may improve with a more condensed battery and with integration into clinical workflows.

**Relevance:** This study provides a practical model for IRFs to adopt and evaluate standardized OM batteries that improve our ability to assess physical function across health systems and settings.

## Introduction

Outcome measure (OM) use in physical therapy has historically been inconsistent with few standardized OM batteries widely adopted into clinical practice.^1–3^ This results in a number of different OMs being used across care setting and healthcare systems. This is problematic because widespread implementation, however, would result in the same OMs being used across settings and systems, facilitating patient-centered care though longitudinal data about an individual’s function^1^ and fostering a shared language among providers to communicate functional status.^1, 4–8^ Further, wide-spread use of standardized OM batteries would generate large-scale data across healthcare systems,^9^ which could be used to evaluate intervention efficacy^1, 10^ and to explore the value of physical therapy.^11, 12^ Unfortunately, reports on processes to implement standardized OM batteries and the success of those efforts remain relatively sparse, representing a critical implementation gap facing rehabilitation.

In neurologic physical therapy, there has been considerable effort to increase and standardize OM usage. For example, the Academy of Neurologic Physical Therapy’s (ANPT) Evaluation Database to Guide Effectiveness (EDGE) task forces evaluated the psychometric properties of OMs for individuals with neurologic conditions.^13–15^ While these task forces stopped short of directly recommending a standardized set of OM (i.e., a standardized OM battery), they were followed by the development of a clinical practice guideline (CPG) that did recommend a standardized OM battery to be used in physical therapy for adults with neurologic conditions across clinical settings and levels of chronicity.^6^ Implementation of this CPG has faced numerous barriers. One barrier is a lack of familiarity and knowledge of OMs by physical therapists (PTs),^1, 16, 17^ though there has been significant effort to generate knowledge translation resources to address this barrier.^18^ Additionally, clinical leadership may not support the implementation of standardized OM batteries^7^ due to concerns about productivity standards and the flow of care.^1^ Although many barriers are similar across diagnosis, clinical setting, and chronicity,^13^ there are unique challenges in inpatient rehabilitation facilities (IRFs), including lack of equipment and space.^19, 20^ Additionally, in the United States, PTs must complete Section GG of the Inpatient Rehabilitation Facility Patient Assessment Instrument (IRF-PAI) at admission and discharge for each patient episode.^21^ While many PTs feel that OMs are important, few are willing to spend more than 20 minutes on assessment.^22^ Since PTs already spend significant time completing Section GG of the IRF-PAI, adding a standard OM battery, like the one suggested by the CPG, likely exacerbates the time constraints present within this setting.^20^

Given the importance of standardized OM batteries and the challenges of implementation within the IRF setting, our purpose was to evaluate the implementation of a standardized OM battery for patients with stroke, brain injury (BI), and SCI in an IRF. This standardized OM battery was based on the CPG and adapted for the IRF setting. We evaluated implementation with Penetration, Sustainability, Acceptability, and Feasibility outcomes as suggested by the Implementation Outcomes Framework.^23^ Insights from this work will inform future implementation of standardized OM batteries in other IRFs, improving our ability to measure physical function across care settings and health systems, and to establish a shared language with other healthcare providers.

## Methods

### Implementation of the Standardized OM Battery at Craig H. Neilsen Rehabilitation Hospital

Craig H. Neilsen Rehabilitation Hospital is a 75-bed, Commission on Accreditation of Rehabilitation Facilities (CARF) accredited IRF with a Center of Excellence Designation in Rehabilitation Services that is affiliated with the University of Utah and primarily treats individuals with neurologic conditions. The implementation effort began in December 2021 with a 45-minute in-person in-service about the standardized OM battery recommended by the CPG (Figure 1). This in-service was delivered to all PTs during work hours (protected, paid time was provided) and resulted in agreement among the PTs and clinical leadership that the standardized OM battery should be implemented to align with best practices.

**Figure 1.**
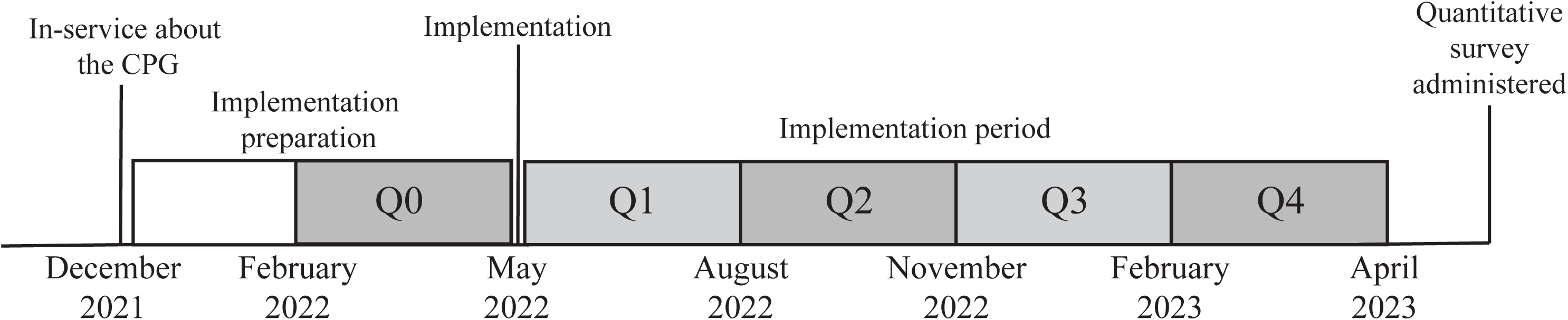
Timeline of implementation of the Transfers, Gait, and Balance Battery. Implementation activities included in-service presentations and a skills lab for physical therapists, as well as integration of the Transfers, Gait, and Balance Battery into the electronic medical record. The quarters are labeled as Quarter 0 (Q0) through Quarter 4 (Q4). Clinical Practice Guideline (CPG)

Preparation for implementation of the standardized OM battery occurred between December 2021 and April 30, 2022 (Figure 1). These efforts were led by the PT supervisor and the IRF director and included staff PTs. First, the team aimed to select the OMs to be implemented, using the CPG as the foundation. The CPG recommends six OMs to assess essential aspects of mobility: 1) the 10 Meter Walk Test (10mWT), which measures walking speed, 2) the 6 Minute Walk Test (6MWT), which measures walking distance, 3) the Berg Balance Scale (BBS), which measures static and dynamic sitting and standing balance, 4) the FGA, which measures walking balance and stair negotiation, 5) the 5 Times Sit to Stand (5xSTS), which quantifies functional lower extremity strength via transfer ability, and 6) the Activities-Specific Balance Confidence Scale (ABC), which measures balance confidence. The team decided to exclude the ABC since the ABC assesses confidence in completing home or community activities, yet individuals admitted to IRFs typically have not experienced home and community activities since acquiring new deficits.^6^ Additionally, the ABC is primarily validated in community-dwelling individuals attending outpatient therapies.^24^ To reflect the omission of the ABC Scale, we refer to the standardized OM battery that was implemented as the Transfers, Gait, and Balance Battery (TGBB).

After finalizing the TGBB, the team partnered with healthcare informaticians to build a TGBB-specific documentation section in the electronic medical record. The documentation section contained all five OMs on the same screen to decrease the number of clicks required when documenting and included individual item descriptions for the BBS and FGA to decrease the need to refer to other resources when documenting the measures. PTs were trained on the execution of the OMs in the TGBB using ANPT online resources.^18^ Training also included two in-services and skills labs which included a post-test to ensure accurate TGBB administration. Participation in the training sessions was mandatory for all PTs, as this effort was considered a quality improvement initiative.

Lastly, during the pre-implementation phase, the expectations for administration of the TGBB were established. Specifically, PTs were expected to administer all five measures of the TGBB within the first three days of admission and again in the three days prior to discharge, resulting in two expected administrations of the TGBB during each episode of care. Additionally, consistent with the CPG, PTs were instructed to document a score of “0” if a patient could not perform the OM due to physical limitations. These expectations were communicated during the initial staff trainings for the TGBB. After the go-live date (May 1, 2022), PTs were updated on group completion rates of the TGBB during quarterly PT-specific staff meetings.

### Implementation outcomes

We assessed four implementation outcomes: Penetration, Sustainability, Acceptability, and Feasibility. These outcomes were identified via the Implementation Outcomes Framework,^23^ which is a taxonomy of conceptually distinct implementation outcomes created to advance understanding of implementation processes. *Penetration* is defined as the number of eligible persons who receive a service, divided by the number of persons eligible for services.^23^ *Sustainability* describes the extent to which a newly implemented practice is maintained within ongoing operations.^23^ *Acceptability* is defined as the extent to which the implemented process is perceived as agreeable, palatable, or satisfactory.^23^ Finally, *Feasibility* is defined as the extent to which an innovation can be successfully used or carried out within an given setting.^23^ We applied the Standards for Reporting Implementation Studies Statement to guide our reporting.^25^

### Data sources

Two data sources were used to quantify our implementation outcomes. We used data from the electronic medical record to quantify Penetration and Sustainability, while data from an electronic survey for PTs at Craig H. Neilsen Rehabilitation Hospital was used to quantify Acceptability and Feasibility. From the electronic medical record, we extracted demographic information (e.g., age, race, sex), clinical information (e.g., length of stay, diagnosis), and the scores on the OMs of the TGBB for each episode of care at admission and discharge. We extracted these data for all episodes of care for three months prior to implementation (February 1, 2022 - April 31, 2022) and for one year after implementation (May 1, 2022 - April 31, 2023). We included all episodes of care, even if an individual had more than one episode of care during the study period, for which the primary diagnosis of stroke, BI, or SCI as determined by Uniform Data System Impairment Categories.^26^ A waiver of consent was approved by the University of Utah Institutional Review Board (IRB #00171405) to obtain these data.

The electronic survey to measure Acceptability and Feasibility was completed as a part of routine project evaluation and, therefore, a waiver of informed consent was granted (IRB #00171405). PTs who were employed either full-time or part-time during the training period and study window were instructed by therapy leadership to complete the survey, which was distributed via Microsoft Teams message in September 2023 (Figure 1). The survey was required for all PTs to complete and included questions about the PT’s clinical background, the impact of TGBB use on clinical practice, and facilitators and barriers to completing the TGBB. The questions relating to practice patterns were based on a previously used survey,^20^ while the questions relating to facilitators and barriers were developed with clinical leadership. All questions were answered using a 5-point Likert scale (1=Completely disagree, 3=Neutral, 5=Completely agree)

### Analysis of Penetration and Sustainability

We used the proportion of opportunities completed to quantify Penetration and Sustainability. To determine the proportion of opportunities completed, we labeled each OM for each individual as “Complete” or “Not Complete.” We considered an OM “Complete” if the PT documented a score, including a 0, in the electronic medical record. We considered a score of 0 as “Complete” because PTs were instructed to assign this score if an individual was not able to complete the OMs. We considered an OM as “Not Complete” if there was no score entered by the PT. For each episode of care, there were two opportunities to complete the OM, one at admission and one at discharge. For our analyses, we treated each opportunity equally and did not differentiate between admission and discharge timepoints. We refer to the proportion of opportunities completed as the “completion rate” throughout the methods and results sections for brevity.

We calculated the completion rate at five timepoints, each representing a 3-month timeframe. The first timeframe was the three months prior to implementation (February 1, 2022 to April 30, 2022) and served as a baseline measurement of OM usage (Figure 1). We refer to this timepoint as Q0. The other four timepoints were consecutive 3-month periods after implementation and are referred to as Q1-Q4 (Figure 1). The completion rate was calculated at each timepoint first for each individual OM and then for the entire TGBB. This was done for the entire cohort, regardless of diagnosis, and for each diagnosis (i.e., stroke, BI, and SCI). For the entire TGBB to be considered complete, all five OMs needed to be completed at the specific administration opportunity (admission or discharge).

We conducted parallel analyses for completion rate of each individual OM and for the entire TGBB. For both, we report the completion rate at each timeframe (Q0-Q4) relative to our target completion rate of 70%, which was based on the median range of PT OM adherence rates in the literature.^7^ Then we quantified Penetration by comparing the completion rate at Q0 and Q1 using a Chi-squared test. Lastly, we quantify Sustainability by comparing the completion rate at Q1 and Q4 with a Chi-squared test. We present the results for each individual OM for the entire cohort and then for each diagnosis. Then we present the results for the entire TGBB for the entire cohort and then for each diagnosis.

When examining the completion rate of the entire TGBB we conducted a secondary analysis to examine the impact of the timing of admission and discharge on completion rates. Specifically, we were interested in determining the impact of transitioning patients between weekday and weekend staff. For this analysis we created two groups. The first consisted of administration opportunities for which the TGBB needed to be done on the weekend. This occurred when admissions occurred on Friday and when discharges occurred on Sunday or Monday. We refer to this group as “Weekend,” while all other opportunities for administration are in the other group, which we call “Weekday.” To assess Penetration and Sustainability in this analysis, we used Chi-squared tests to compare Q0 to Q1 and Q1 to Q4, respectively, within the Weekend and Weekday groups. Fisher’s exact tests were used when sample sizes were small or expected cell counts were less than five. We conducted this analysis in the entire cohort and for each diagnosis.

### Analysis for Acceptability and Feasibility

We assessed Acceptability and Feasibility with use of the electronic survey administered to the PTs. We present descriptive information regarding each survey question, and when presenting the results we combined the “Completely Disagree” and “Somewhat Disagree” responses into a “Disagree” category and the “Somewhat Agree” and “Completely Agree” responses into an “Agree” category. We used questions about practice changes to measure acceptability, while questions about facilitators and barriers measured feasibility. All survey questions are provided in Supplemental materials (Survey Questions). All analyses were completed in R (R v.4.4.2; R Core Team) with an alpha of 0.05. The funders played no role in the design, conduct, or reporting of this study.

## Results

### Penetration and Sustainability of Individual OMs within the TGBB

This analysis included 773 episodes of care from 702 unique patients between February 1, 2022 - April 30, 2023. Details about those episodes of care and individuals included are in Table S1. First, we examined the completion rate for each of the OMs within the TGBB within the full cohort. All five OMs demonstrated an increase in the completion rate from Q0 to Q4, with all measures exceeding the 70% threshold by Q4 (Figure 2a). BBS was the first OM to exceed to 70% threshold, with this OM being completed during 72.4% (265/366) of the opportunities in Q1. Chi-squared tests assessing Penetration showed a significant increase in the completion rate from Q0 to Q1 for all OMs (all p<0.001). For example, the FGA increased from 29.9% (73/244) of opportunities completed in Q0 to 63.9% (234/366) at Q1. When examining Sustainment in the full cohort, all five OMs demonstrated a significant increase in completion rate from Q1 to Q4 (Table 1; p<0.05 for all). For example, the BBS completion rate increased from 72.4% (265/366) at Q1 to 80.4% (239/297) at Q4. This significant result indicates that there was continued improvement in the completion rate during the year following implementation.

**Figure 2.**
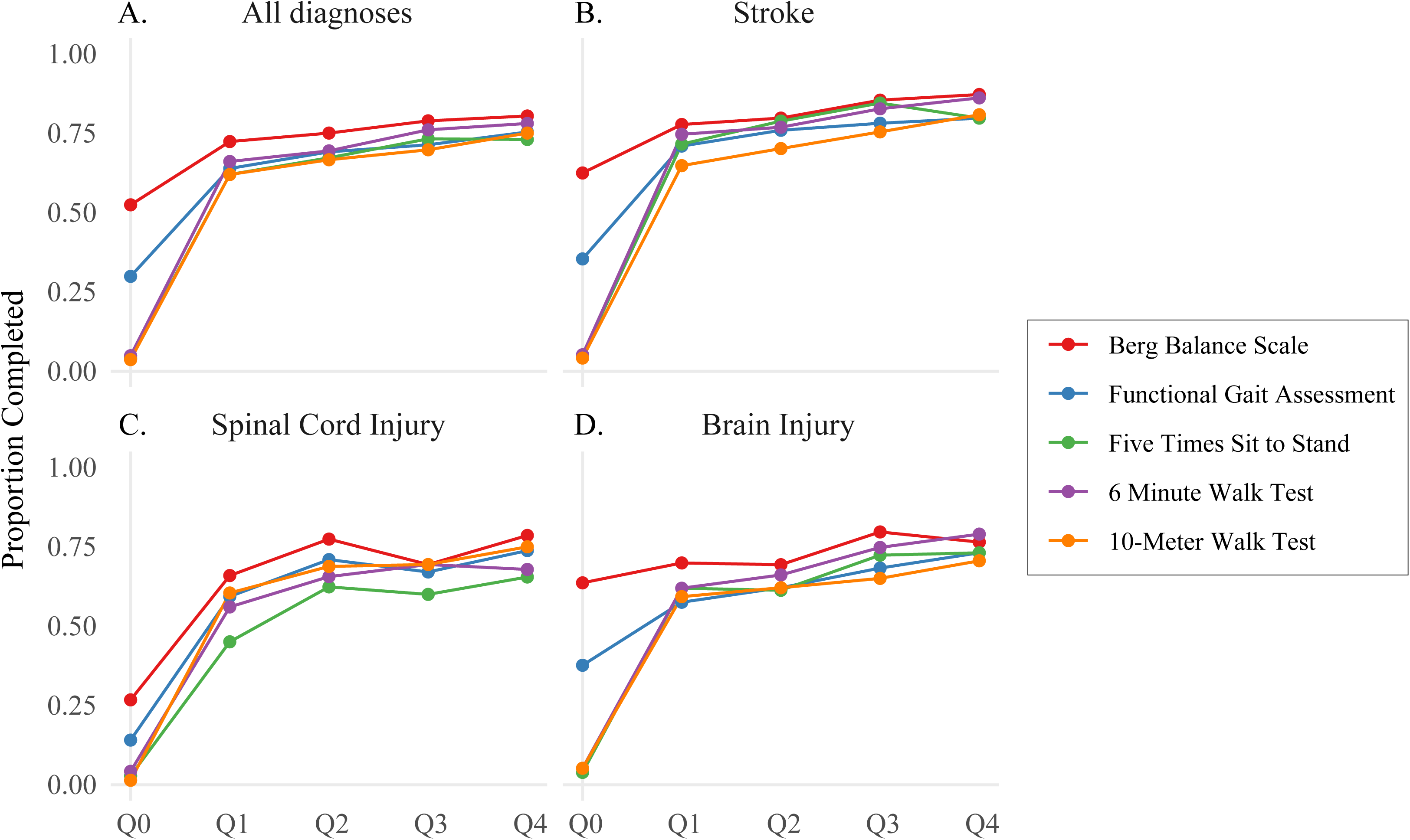
Completion rates of individual outcome measures within the Transfers, Gait, and Balance Battery for all diagnoses (a) and individual diagnoses (b-d).

**Table 1.**
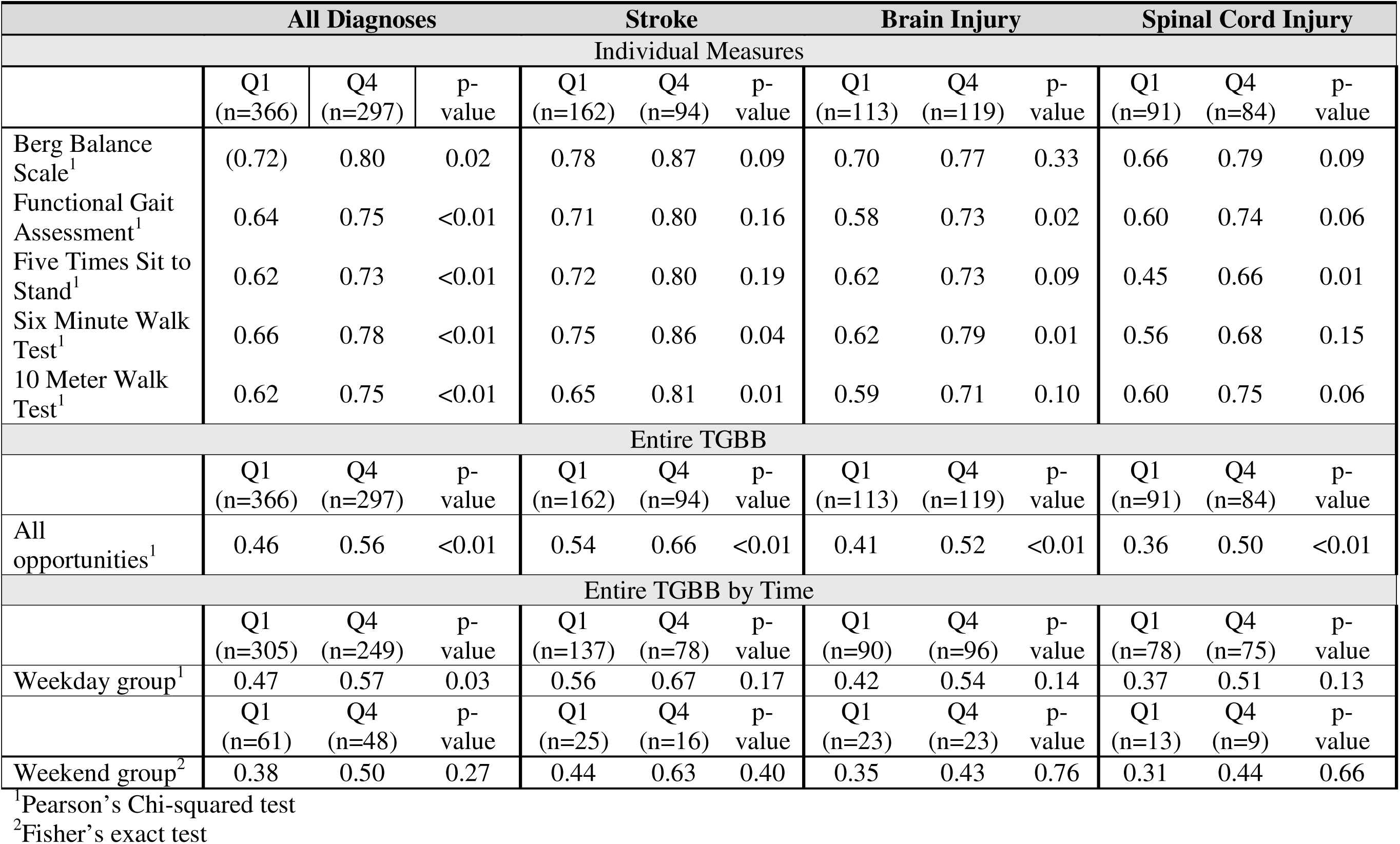
Comparison of completion rates at Quarter 1 and Quarter 4 for each outcome measure by diagnosis.

**Table 2.**
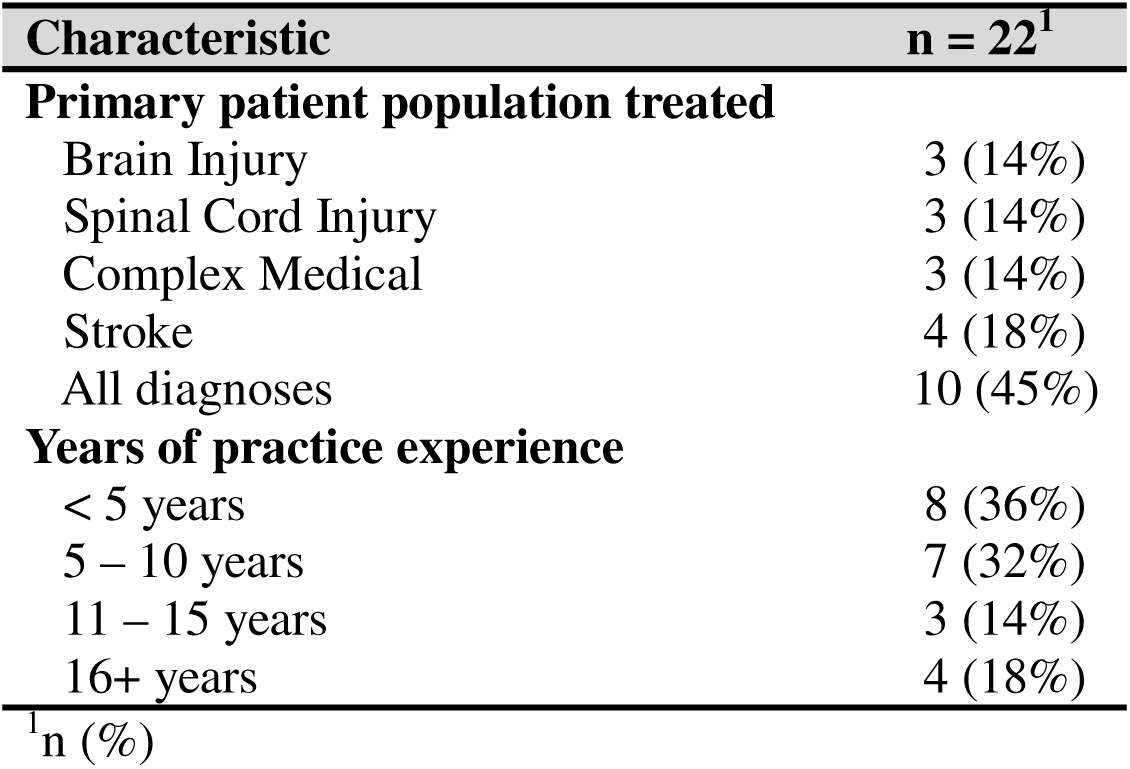
Physical Therapist Characteristics.

| Table 2. Physical Therapist Characteristics |  |
| --- | --- |
| Characteristic | n = 22<sup>1</sup> |
| **Primary patient population treated** |  |
| Brain Injury | 3 (14%) |
| Spinal Cord Injury | 3 (14%) |
| Complex Medical | 3 (14%) |
| Stroke | 4 (18%) |
| All diagnoses | 10 (45%) |
| **Years of practice experience** |  |
| < 5 years | 8 (36%) |
| 5 – 10 years | 7 (32%) |
| 11 – 15 years | 3 (14%) |
| 16+ years | 4 (18%) |
| <sup>1</sup>n (%) |  |

When evaluating the completion rate for each OM by diagnosis, all OMs exceeded the 70% threshold for each diagnosis by Q4 except for the 5xSTS and 6MWT for the SCI diagnosis, which remained below the 70% threshold throughout all timepoints (Figure 2b-d). With the exception of the BBS for individuals with BI, Chi-squared tests for Penetration showed a significant increase in the completion rate from Q0 to Q1 for all OMs within diagnosis (p<0.05 for all). The results for Sustainability for the specific diagnoses were not as uniform, with some OMs demonstrating continued improvement and others not demonstrating this improvement from Q1 to Q4 (Table 1). For example, for individuals with CVA, the 6MWT and 10mWT demonstrated a significant increase in completion rate between Q1 and Q4 (p<0.05). There was no significant difference in completion rate for BBS, FGA, and 5xSTS in the CVA diagnosis; however, the completion rate did not decrease.

### Penetration and Sustainability of the entire TGBB

Next, we examined the completion rate for the TGBB in its entirety. For the full cohort, 45.6% of opportunities had the entire TGBB completed during Q1. Although the completion rate for the entire TGBB increased over the year after implementation, it never exceeded the 70% goal. (Figure 3a; purple line). The assessment of Penetration demonstrated a significantly higher proportion of opportunities when the entire TGBB was completed for Q1 compared to Q0 (p<0.001 for all), indicating that although the 70% threshold was not met, implementation resulted in a significantly increased administration of the entire TGBB. The Sustainability assessment demonstrated a significantly higher completion rate for Q4 compared to Q1 in the full cohort (p<0.01), indicating continued increases in the completion rate of the entire TGBB over the year after implementation.

**Figure 3.**
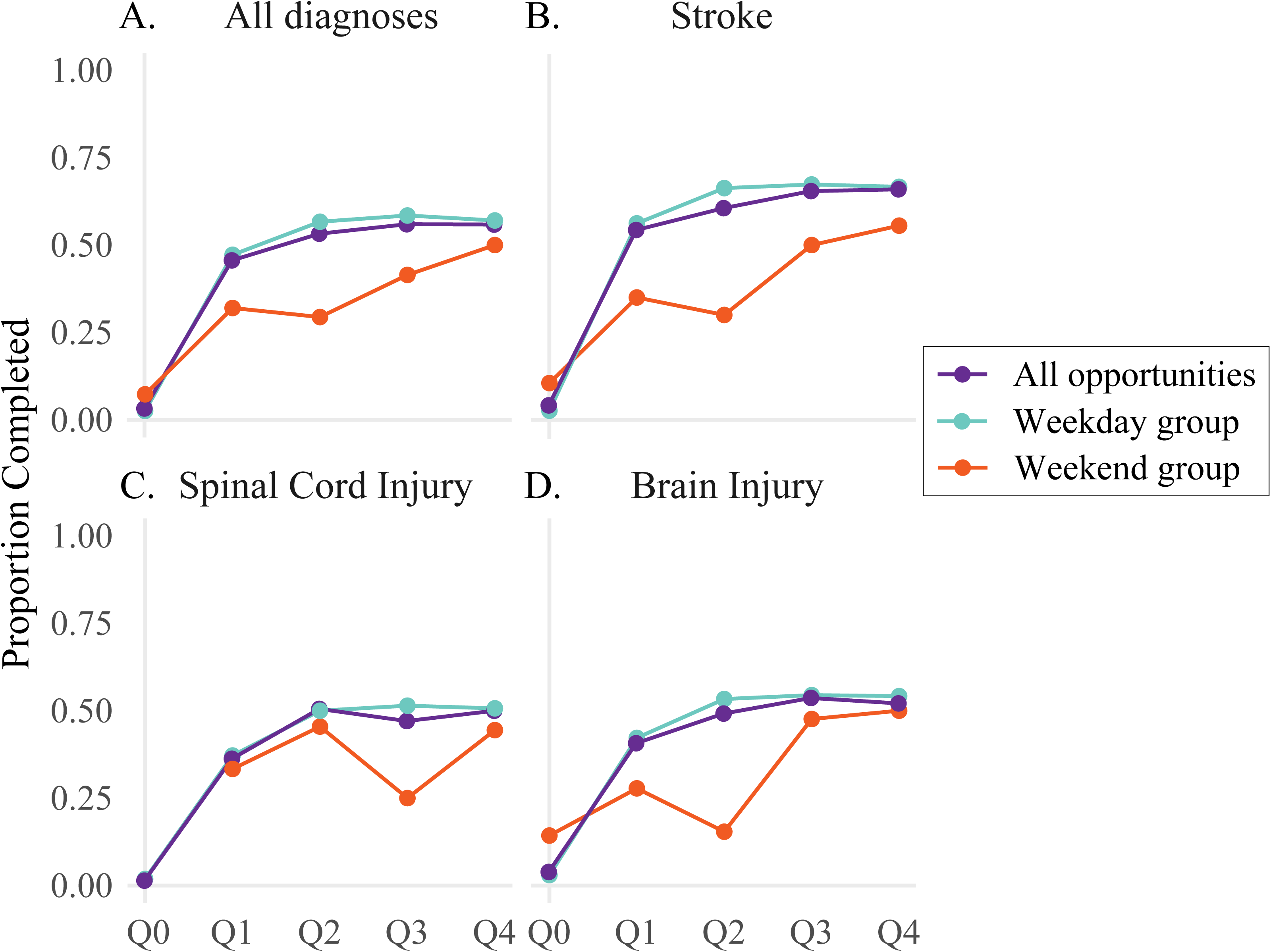
Completion rate of the Transfers, Gait, and Balance Battery for all diagnoses (a) and individual diagnoses (b-d). The Weekend group represents the administration opportunities with an admission on Friday or discharge on Sunday or Monday; the Weekday group represents all other opportunities; All Opportunities represents all opportunities for administration regardless of admission and discharge timing. The Spinal Cord Injury diagnosis had no weekend administration opportunities in Q0.

When evaluating the completion rate by diagnosis, we also observed that the completion rate did not exceed the 70% goal for any of the three diagnoses (Figure 3b-d, purple line). Despite this, all three diagnoses demonstrated Penetration as evident by an increase in completion rate from Q0 to Q1 (p<0.001 for all), though the SCI diagnosis demonstrated the lowest completion rate of 36.3% (Figure 3b-d). For the three diagnoses, there was a trend of increasing completion rate from Q1 to Q4, however, this did not reach significance (p>0.09 for all). For example, the CVA diagnosis had a 54.3% (88/162) completion rate in Q1 and a 65.9% (62/94) completion rate in Q4 (Table 1).

In our secondary analysis, we examined the impact of the timing of admission or discharge on the completion rate for the entire TGBB. When examining Penetration, both the Weekend and Weekday groups demonstrated a significantly higher completion rate for Q1 compared to Q0 (p<0.01; Figure 3a, teal and orange), though the Weekday group demonstrated a higher completion rate of 47.2% (144/305) at Q1 compared to a 37.7% (23/61) completion rate for the Weekend group (Table 1; Figure 3a, teal and orange). When examining Penetration for individual diagnoses the patterns were similar, such that there was a significant increase in completion rate for both the Weekend and Weekday groups for individuals with stroke and SCI and for the Weekday group for BI (p<0.05; Figure 3b-d). The change from Q0 to Q1 was not significant for the Weekend group in BI diagnosis (p=0.22). When examining Sustainability, both the Weekend and Weekday groups demonstrated an increase in completion rate from Q1 to Q4, though only the Weekday group demonstrated a significant increase (p=0.03). The completion rates increased from Q1 to Q4 in both the Weekday and Weekend groups within each diagnosis, though none of the increases were statistically significant (Table 1).

### Acceptability and Feasibility of the TGBB

All eligible staff PTs (22/22) completed the survey. Fifteen PTs had less than 10 years of experience (Table 1). Three PTs were board-certified specialists in Neurologic Physical Therapy, though all PTs surveyed had extensive experience treating individuals with neurologic conditions.

Regarding Acceptability of the TGBB, PTs generally agreed that the implementation of the TGBB resulted in positive practice changes (Figure 4a). For example, a majority of PTs felt that the TGBB supported a “Culture of data” [90.9% (20/22) “Agree”] and increased the use of OMs in clinical decisions [77.3% (17/22) “Agree”].

**Figure 4.**
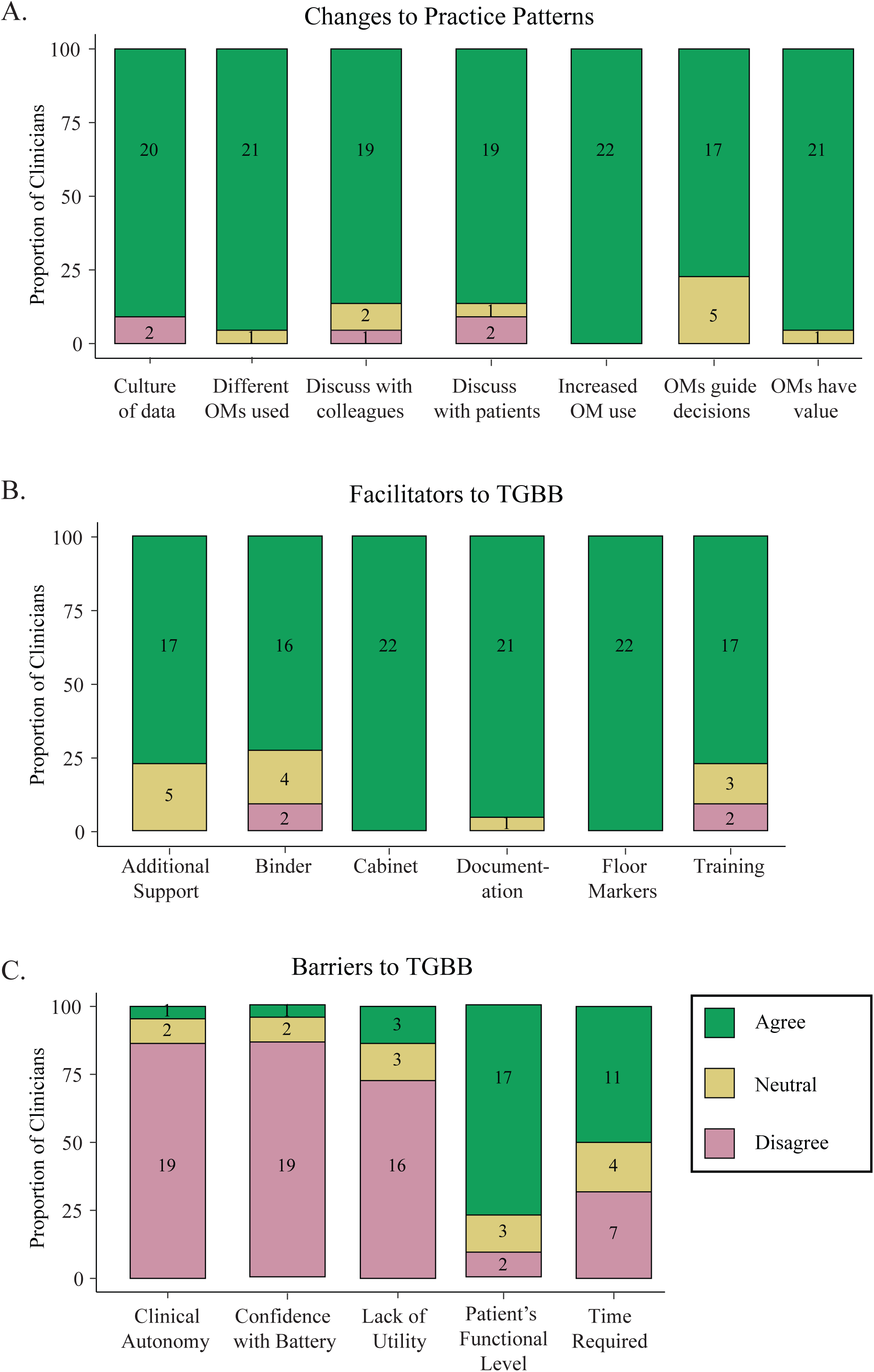
Results of physical therapist survey for a) Acceptability of the Transfers, Gait, and Balance Battery and b) facilitators and c) barriers to using the Transfers, Gait, and Balance Battery.

The second part of the survey asked about the systems that impacted the Feasibility of performing the TGBB (Figures 4b-c). PTs found the “Outcomes cabinet,” which contained all necessary materials to perform the TGBB, permanent lines on the gym floor to demarcate distances for each test, and a specific TGBB documentation section most helpful. Finally, PTs also endorsed several barriers to completing the TGBB (Figure 4c). The most widely endorsed barriers were a patient’s functional level [77.3% (17/22) “Agree”] and the time required to complete the OMs [50% (11/22) “Agree”].

## Discussion

This work describes the implementation of the TGBB, a standardized OM battery based on a CPG recommending OMs for individuals with neurologic diagnoses.^6^ We used electronic health record data from Craig H. Neilsen Rehabilitation Hospital over a 15 month period to evaluate Penetration and Sustainability and an electronic survey of PTs to evaluate acceptance and feasibility. While the completion rate for each individual OM exceeded our target threshold of 70%, the completion of all OMs in the TGBB did not exceed that threshold. However, there was a significant increase in the completion rate of the full TGBB from pre-implementation to the first 3 months after implementation. PTs endorsed a positive impact of the TGBB on clinical practice, demonstrating acceptance, and identified concrete items that improved or hindered the feasibility of completing the TGBB.

When analyzing Penetration, we observed high completion rates for single OMs but lower rates for the entire battery. This indicates that PTs seem to prioritize specific OMs rather than completing the entire battery. Based on responses to the survey, the prioritization could be based on a patient’s functional level. Since the CPG recommends that OMs in the standardized OM battery only be scored if the patient has goals and the capacity to improve in that area,^6^ some of these clinical decisions may have aligned with best practices but not with the established protocol; however, we were unable to determine this based on the data documented within the electronic medical record. Additionally, there is variation in the amount of time required to complete different OMs within the battery. It is possible that PTs selected OMs to complete based on time constraints within a session. This is supported by a majority of PTs endorsing time as a barrier to completing the TGBB. Based on the Penetration rate of the full TGBB, an OM battery consisting of five separate OMs might have been too challenging to complete in this clinical setting. Despite this, the TGBB demonstrated excellent Sustainability throughout the year long period, with no decline observed in any of the analyses.

The timing of admission and discharge could provide an additional explanation for the higher completion rates for individual OMs but lower rates for the entire battery. Though the Weekday and Weekend groups demonstrated similar overall trajectories of completion rate from Q0 to Q4, the completion rates themselves were different, with the Weekend groups having lower completion rates than the Weekday group at all timepoints after implementation (Q1-Q4). This suggests that the transition between weekday and weekend staffing could affect completion rates due to the added step of coordinating and communicating which OMs need to be completed between two different PTs. Additionally, patients receive fewer physical therapy minutes during the weekend compared to the weekday, which could contribute to increased challenge for administering the full battery. These potential explanations provide an additional barrier to completion of a standardized OM battery in the IRF setting.

When analyzing the Penetration and Sustainability of the individual OMs, the BBS was the first OM to reach the 70% completion rate threshold and had the highest completion rate during the pre-implementation phase. PTs may have been more familiar with administering the BBS, resulting in the highest completion rates compared to the other OMs. Additionally, the BBS does not have items requiring ambulation, therefore it may have been more appropriate for a wider range of patients to complete. The 6MWT had one of the lowest completion rates during the pre-implementation phase and then increased to the second highest completion rate by Q3. These patterns suggest that clinician familiarity can influence OM completion, reinforcing the need for specific training and protocols to improve battery completion rates.

All three diagnoses demonstrated a similar trajectory of increase in completion rate for the full TGBB from Q0 to Q4, though the CVA diagnosis achieved and sustained a higher completion rate compared to the BI and SCI diagnosis. The trajectories were different between diagnoses when examining the individual OMs. For example, the SCI diagnosis demonstrated lower completion rates for all OMs in the pre-implementation phase. This could potentially be attributed to the functional level of individuals with SCI in the IRF setting, which would align with the survey finding that a barrier to completion of the OM battery was a patient’s functional level. Additionally, the 5xSTS test had the lowest completion rate over time in the SCI diagnosis, while the 10mWT had the lowest completion rate in the CVA and BI diagnoses. This variability indicates that further analysis is warranted to determine if characteristics of the individual OMs made them more or less appropriate for specific diagnostic groups in this clinical setting.

The quantitative survey revealed changes in practice patterns that demonstrated acceptability of the battery as well as identifying barriers and facilitators to implementing this CPG in the IRF setting. First, PTs indicated that implementing the TGBB had a positive effect on their practice by creating a culture of data and increasing OM use to guide clinical decision making. This shift in practice patterns supports an emphasis on patient-centered care and improved infrastructure to track functional outcomes across the continuum of care.

The survey revealed tangible information regarding the feasibility of TGBB implementation. One of the primary barriers to completing the OM battery was a patient’s functional level. This finding aligns with previously identified barriers to OM use, such as challenges with the compatibility of the OMs in the IRF setting.^19^ Additional research is needed to understand the impact of function on completion of a standardized OM battery based on the Core Set of Outcome Measures CPG.

Environmental support, such as cabinets with TGBB supplies and permanent floor markings to demarcate distance, facilitated TGBB completion. This aligns with previous literature identifying space and equipment limitations as barriers to standardized OM use in the IRF setting.^19, 20^ Our work further emphasizes the importance of environmental support for OM implementation in IRFs.

### Limitations

This work provides valuable insights into implementing a standardized OM battery in an IRF, yet it has limitations. First, it was not possible to identify why individual OMs for individual patients were not completed based on the data that were collected. The CPG on which the standardized OM battery is based suggests administering the recommended OMs as long as a patient has goals and the capacity to improve in the specific construct that the given OM assesses.^6^ We assumed that most individuals had goals and the capacity to improve in these mobility domains; however, we were not able to confirm this for every episode. In situations where there were no goals or ability to improve in a specific domain, it might have been appropriate to not complete certain measures, but we were unable to determine this. Additionally, our work could be strengthened with further exploration of the barriers to implementation identified by PTs, such as quantifying the time required for OM battery completion to better understand how long PTs are willing to spend on OM administration in this setting and if that explains non-compliance with the protocol. Similarly, further exploration of the relationship between a patient’s functional level and PTs’ administration of OMs would provide further insights as to the completion of the standardized OM battery in this clinical setting. Lastly, this project was completed at a single site; including additional IRF locations could provide a broader understanding of the TGBB’s use in this care setting and practice patterns.

### Conclusions

This work demonstrates that a standardized OM battery based a CPG recommending OMs in individuals with neurologic diagnoses^6^ can be implemented in the IRF setting, though implementation of fewer OMs might have a higher completion rate than a five OM battery in this setting. Patients’ functional level and the design of the electronic medical record can influence PTs’ ability to complete the battery. Future research should analyze the OM scores to examine the change in function from IRF admission to discharge and assess the success of standardized balance and mobility OMs with fewer floor effects. This work can serve as a model for IRFs to implement standardized OM batteries, improving our ability to capture metrics of physical function across health systems.

## Data Availability

All data produced in the present study are available upon reasonable request to the authors

## Notes

The authors declare no conflict of interest.

### Competing Interest Statement

The authors have declared no competing interest.

### Funding Statement

This study did not receive any funding

### Author Declarations

A waiver of consent was approved by the University of Utah Institutional Review Board (IRB #00171405) to obtain these data.

