## Supplemental Materials for "Implementation of a Gait and Balance Battery of Outcome Measures in an Inpatient Rehabilitation Hospital"

| Table S1. Demographic and Clinical Characteristics | | | | | |
| --- | --- | --- | --- | --- | --- |
|  | **Overall^1^** | **Brain Injury^1^** | **Spinal Cord Injury ^1^** | **Stroke^1^** | **p-value**^2^ |
| **Person-level variables** | | | | | |
| n | 702 | 246 | 189 | 267 |  |
| Race |  |  |  |  | 0.3 |
| Non-White | 79 (11%) | 22 (8.9%) | 20 (11%) | 37 (14%) |  |
| Unknown | 64 (9.1%) | 21 (8.5%) | 22 (12%) | 21 (7.9%) |  |
| White | 559 (80%) | 203 (83%) | 147 (78%) | 209 (78%) |  |
| Gender |  |  |  |  | 0.5 |
| Female | 279 (40%) | 102 (41%) | 68 (36%) | 109 (41%) |  |
| Male | 423 (60%) | 144 (59%) | 121 (64%) | 158 (59%) |  |
| **Episode-level variables** | | | | | |
| n | 773 | 278 | 212 | 283 |  |
| Age at Admission | 58.5 (18.2) | 55.2 (20.0) | 56.0 (18.6) | 63.8 (14.6) | <0.001 |
| Length of Stay (days) | 18.8 (12.7) | 16.3 (10.6) | 24.8 (17.6) | 16.7 (7.9) | <0.001 |
| Discharge Location |  |  |  |  | 0.075 |
| Facility | 218 (28%) | 92 (33%) | 53 (25%) | 73 (26%) |  |
| Home | 555 (72%) | 186 (67%) | 159 (75%) | 210 (74%) |  |
| ^1^Mean (SD); n (%) | | | | | |
| ^2^Kruskal-Wallis rank sum test; Pearson's Chi-squared test | | | | | |

**Clinical Staff Survey about TGBB**

A. Please answer the following questions about whether the implementation of the TGBB has changed your practice using the following scale: 1) completely disagree, 2) somewhat disagree, 3) neutral (neither agree nor disagree), 4) somewhat agree, or 5) completely agree.

After TGBB initiation (if you were not here prior to initiation of the TGBB, answer these in relation to prior to joining the IRF):

1. I have increased the use of outcome measures in my clinical practice.
2. I have changed which outcome measures I use in my clinical practice.
3. I use the outcome measure results to guide my clinical decision making.
4. I have more discussions with my patients about their outcome measurement results.
5. I have more discussions with colleagues about outcome measure results.
6. The culture in our department has shifted to discuss patient-related data (i.e. outcome measure results) instead of patient observations (i.e. patient walks slowly, has poor balance, etc).
7. I better understand the value outcome measures add to clinical practice.
B. The following tool/resource made it easier and more time efficient for me to complete the TGBB: 1) completely disagree, 2) somewhat disagree, 3) neutral (neither agree nor disagree), 4) somewhat agree, or 5) completely agree.
   1. “Outcomes cabinet” with items including cones, slipper, step stool, etc
   2. Binder containing instructions and score sheets for each outcome measure
   3. Permanent lines on floor demarcating distance for each outcome measure
   4. Epic section with all required outcome measures listed together for documentation
   5. Training/in-service with skills day lab prior to implementation
   6. Additional support from rehab aides or students
C. The following factors impair my ability to complete the TGBB: 1) completely disagree, 2) somewhat disagree, 3) neutral (neither agree nor disagree), 4) somewhat agree, or 5) completely agree.
1. The time required
2. My patient’s ability
3. Comfort with the outcome measures
4. Limiting my clinical autonomy/clinical reasoning skills
5. Lack of clinical utility
